# An Efficient and Interpretable Learning Approach for Large-Scale Histopathology Data

**DOI:** 10.64898/2026.04.30.26352196

**Authors:** Charles Moore, Vibhuti Gupta, Himanshu Tripathi, Subash Neupane

## Abstract

Prostate cancer (PCa) remains one of the leading causes of cancer-related mortality among men, and histopathological analysis of prostate biopsy specimens is central to diagnosis and risk stratification. Whole-slide Images (WSIs) capture rich morphological information, but their gigapixel scale and the large number of extracted tissue patches make exhaustive annotation and model training computationally expensive. Attention-based Multiple Instance Learning (MIL) has emerged as an effective weakly supervised framework for WSI analysis, enabling slide-level prediction without requiring patch-level annotations. However, training MIL models on large histopathology cohorts remains resource intensive because many extracted patches are non-informative, and some patches are often processed repeatedly during training. To address these challenges, we propose an efficient and interpretable learning framework for large-scale histopathology analysis. Our method combines a pathology-pretrained UNI encoder, a Clustering-constrained Attention Multiple instance learning-Single Branch (CLAM-SB) attention-based MIL model, and a window-based training strategy that reduces computational overhead while preserving predictive performance. The paper illustrates our proposed approach and experiments on TCGA-PRAD WSIs for the PCa patients. Processing 189,600 sampled patches across 79 WSIs with our proposed approach reduced total training time by 57.5% (20 to 8.5 hours for 5 epochs) and 41.4% (27 to 16 hours for 10 epochs), respectively, underscoring its potential as a practical and resource-efficient strategy for scalable prostate histopathology analysis.

## I. Introduction

Prostate cancer (PCa) is one of the leading causes of cancer-related death among men [1, 2]. In the United States alone, approximately 313,780 new cases and 35,770 deaths were projected for 2025 [3]. Around 80% of PCa cases occur in men aged 70 or older, with the survival rates approaching 100% for early-stage diagnosis but declining to 38% for distant-stage PCa [3]. The burden of disease is not uniformly distributed across populations, and African American men experience a substantially higher incidence (67%) of PCa than Caucasian American men [4, 5].

PCa risk diagnostics and stratification involve multiple steps, including Prostate-Specific Antigen (PSA) test, digital rectal exam, biopsy, and Gleason grading [6]. Among these, histopathological examination of prostate biopsy specimens remains the clinical gold standard for PCa diagnosis because it provides critical morphological evidence for tumor detection, grading, and risk stratification. However, histopathology WSIs are extremely large, often containing millions of pixels and thousands of tissue patches, which creates substantial computational challenges for downstream analysis. A single WSI can contain gigapixel-level information, often requiring several gigabytes of storage, making direct end-to-end processing infeasible with standard deep learning architectures. As a result, histopathology WSIs are divided into smaller tiles or patches, which significantly increases data volume, memory requirements, and training time. In addition, only a subset of extracted patches is diagnostically informative, while many correspond to background or non-informative tissue regions. Variability in staining, tissue preparation, and scanner artifacts further complicates robust analysis by introducing noise and domain shifts. Together, these factors motivate the development of efficient and scalable learning strategies for large-scale WSI analysis.

Multiple Instance Learning (MIL), particularly attention-based variants, has become a widely used weakly supervised framework for WSI analysis because it supports slide-level prediction without the need for path-level annotations [7, 8]. Recent studies in prostate histopathology have explored MIL-based strategies that improve feature interaction, patch representation [9], and domain-specific aggregation [10, 11]. Despite these advances, training attention-based MIL models remains computationally expensive at scale. Each slide may contain thousands to hundreds of thousands of patches, requiring repeated feature extraction and attention-based aggregation over large bags of instances. Because many extracted patches are non-informative, substantial computation is spent on regions that contribute little to the final prediction. This leads to high memory consumption, slow convergence, and limited scalability, particularly in resource-constrained training environments. For example, a WSI with 50,000 patches and 1,024-dimensional embeddings requires about 200 MB of intermediate feature storage, and scaling this across hundreds of slides per epoch can quickly exhaust GPU memory.

To address these challenges, we propose an efficient and interpretable learning approach for large-scale histopathology analysis. Our method combines a pathology-pretrained patch encoder with an attention-based MIL architecture and a window-based training strategy designed to reduce computational overhead while preserving predictive performance. We evaluate the proposed approach on TCGA-PRAD [12] and analyze both predictive accuracy and computational efficiency across multiple training configurations. The main contributions of this paper are as follows:

1. We propose a compute-efficient and interpretable training framework for large-scale prostate histopathology analysis that combines UNI-based patch encoding, CLAM-SB attention-based aggregation, and a window-based training strategy.
2. We perform an empirical evaluation on TCGA-PRAD using accuracy, ROC-AUC, precision, recall, F1-score, and training time to characterize the trade-off between predictive performance and computational efficiency.
3. We show that competitive slide-level prostate cancer risk classification can be achieved without processing all extracted patches at every training stage, highlighting a practical strategy for resource-constrained pathology learning.

## II. Related Work

Recent advances in computational pathology have shown that deep learning on WSIs can improve PCa risk prediction and prognostication. Several studies have moved beyond traditional Gleason grading to directly model clinically meaningful outcomes such as biochemical recurrence and disease progression [13–15]. Methodologically, weakly supervised learning approaches, particularly Multiple Instance Learning (MIL) and graph-based aggregation frameworks, have enabled scalable WSI analysis using only slide-level labels, achieving high diagnostic and grading performance across large cohorts [16, 17]. Additional studies have demonstrated that deep learning models can accurately predict Gleason grades directly from WSIs without region-level annotations, serving as strong proxies for disease aggressiveness [18]. Collectively, these works establish WSIs as a powerful modality for PCa risk modeling and motivate the development of more robust and multimodal approaches for improved prognostic prediction.

Recently there are some works on efficient processing of WSIs through weakly supervised learning, patch selection, and efficient representation learning [8, 19–24]. [8] developed an attention-based Multiple Instance Learning framework, such as CLAM that enables scalable WSI analysis by focusing only on diagnostically relevant regions using slide-level labels, significantly reducing annotation and computation requirements. Prototype-based approaches such as ReMix further improve efficiency by replacing large patch sets with representative cluster centroids, thereby reducing memory and training time [19]. Similarly, clustering-based feature aggregation methods provide compact slide-level representations while minimizing computational overhead [20]. More recent approaches such as hierarchical distillation MIL selectively filter informative regions, reducing inference cost while maintaining accuracy [21], while hard instance mining strategies focus learning on the most informative patches to improve both efficiency and generalization [22]. In addition, unsupervised methods such as SAMPLER demonstrate that efficient statistical representations can achieve competitive performance with significantly reduced training time [23]. Finally, system-level innovations such as on-the-fly patch extraction eliminate the need for expensive preprocessing pipelines, further improving scalability [24]. Overall, these approaches highlight a shift toward scalable and resource-efficient WSI modeling, enabling practical deployment of deep learning in computational pathology.

Unlike prior approaches that focus on reducing data dimensionality or selecting informative regions, our window-based training strategy introduces a fundamentally different paradigm by optimizing the training process itself. By sequentially exposing subsets of the dataset across epochs, our approach significantly reduces computational burden while preserving full data fidelity, making it complementary to existing MIL, sampling, and representation learning methods.

## III. Method

### A. Dataset and Preprocessing

Whole-Slide histopathology Images (WSIs) present substantial computational challenges because of their extremely large spatial resolution, often exceeding gigapixel scale. Directly loading and processing full-resolution WSIs is infeasible in most deep learning workflows. To address this challenge, we adopted a multi-stage preprocessing and data loading pipeline that converts WSIs into manageable patch-level inputs for model training. The overall preprocessing workflow consisted of (1) slide preprocessing and patch extraction using TRIDENT [25], (2) hierarchical dataset organization, and (3) dynamic patch-level loading during model training.

We utilized WSIs from the TCGA-PRAD [12] cohort. After excluding cases with unusable whole-slide images or insufficient clinical annotations, 243 patients were retained for analysis. Each patient was associated with one or more diagnostic WSIs acquired from biopsy or surgical specimens. These slides capture clinically relevant morphological features, including glandular architecture, stromal composition, and cellular organization, which are important for PCa diagnosis and risk assessment.

All slides were preprocessed using the TRIDENT framework, which performs tissue segmentation, removes background regions such as whitespace and non-tissue areas, and extracts fixed-size image patches from tissue-containing regions. In our implementation, TRIDENT generated patches of size 256 × 256 pixels. This patch-based representation converts each WSI into a set of smaller image regions that can be processed efficiently while preserving local morphological information. During training, valid patches were loaded dynamically from slide-specific directories. To maintain balanced computational workloads across slides and reduce unnecessary processing, we randomly sampled up to 2,400 patches per slide during each training iteration, resulting in a maximum of 189,600 sampled patches per iteration in our implementation. This sampling strategy improves efficiency while preserving exposure to diagnostically relevant tissue regions.

### B. Our Proposed Framework

Our framework combines a pathology-pretrained patch encoder, an attention-based MIL model, and a compute-efficient window-based training strategy as shown in Figure 1. Specifically, UNI [26] is used to generate patch-level embeddings, CLAM-SB is used to aggregate patch representations and produce slide-level predictions, and the proposed window-based training schedule is designed to reduce the computational burden of training on large WSI cohorts. Below are the details on the WSIs processing steps:

**Fig. 1.**
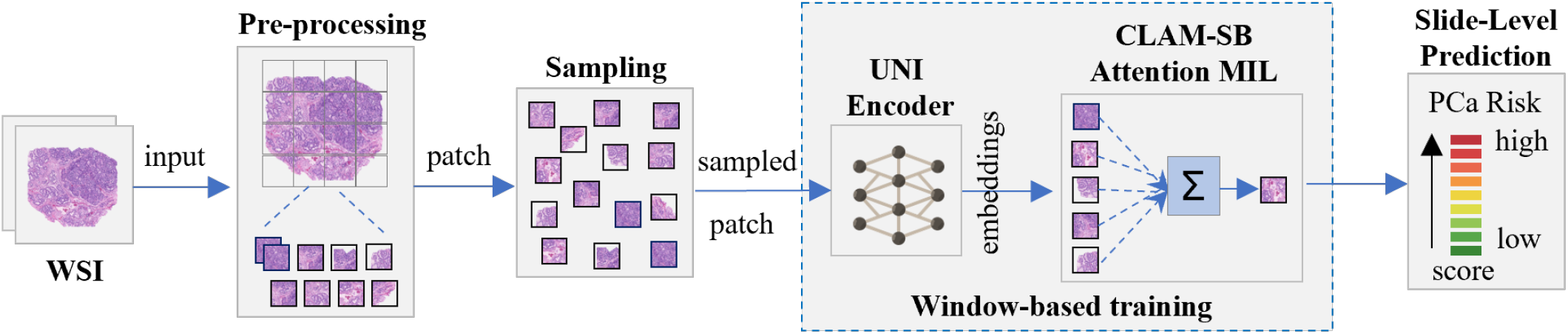
Overview of the proposed framework. Whole-slide histopathology images are preprocessed using TRIDENT for tissue segmentation, background removal, and patch extraction. Sampled patches are encoded using the pathology-pretrained UNI model and aggregated with CLAM-SB through attention-based multiple instance learning to generate slide-level predictions.

#### 1) Patch Feature Extraction (UNI Encoder)

Given a bag of patches *B* = {*x*_1_, …, *x*_*N*_} from a WSI, we extract patch embeddings using UNI:

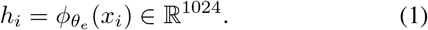

where 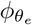 denotes the UNI feature extractor, and *h*_i_ is the resulting 1024-dimensional representation of patch *x*_*i*_. These embeddings provide compact pathology-informed representations for downstream slide-level classification.

#### 2) Attention-Based MIL Aggregation (CLAM-SB)

Given embeddings *H* = {*h*_1_, …, *h*_*N*_}, CLAM-SB computes gated attention weights over instances as follows:

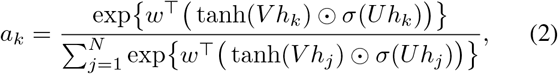

The attention weights are then used to form a slide-level representation through attention pooling:

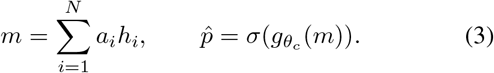

where, *m* denotes the aggregated slide representation, and 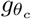 is the slide-level prediction head. This attention-based formulation enables the model to emphasize the most informative tissue regions while preserving interpretability through patch-level attention scores.

#### 3) Instance-Level Supervision and Training Objective

In addition to the bag-level classification objective, we adopt CLAM’s auxiliary instance-level supervision strategy. Specifically, the model selects the top-*k* most-attended and bottom-*k* least-attended patches, with *k* = 8, and uses them to compute an auxiliary instance loss ℒ_instance_. The total training loss is defined as:

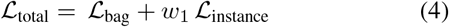

This additional supervision improves patch-level discrimination while supporting robust slide-level prediction.

#### 4) Window-Based Training Strategy

Training deep learning models on WSI datasets is computationally expensive due to the large number of patches generated from each slide. To improve training efficiency, we partition the training data into equal-sized windows containing patient subsets, rather than training on all available data simultaneously. This approach reduces memory overhead and shortens training time while still enabling the model to learn gradually over multiple training iterations.

Let 𝒫= {*p*_1_, *p*_2_, …, *p*_*N*_} denote the full set of training patients. At the beginning of training, patient indices are randomly shuffled to produce a permuted sequence 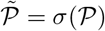, where *σ* is a random permutation. The shuffled set is then partitioned into *W* non-overlapping windows:

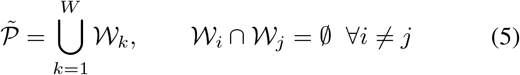

where each window *𝒲*_*k*_ is defined as

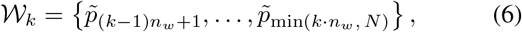

with *n*_*w*_ denoting the number of patients per window and *W* = ⌈*N*/*n*_*w*_⌉ denoting the total number of windows.

For each window, all WSIs associated with the patients in that subset are processed. Patch embeddings are extracted using the frozen UNI encoder, aggregated using CLAM-SB, and used to optimize the slide-level classification objective. After training on one window, the learned model weights are frozen and fine-tuned on each subsequent window. This staged training strategy reduces memory usage, disk I/O overhead, and repeated computation while still allowing the model to learn from the full cohort over time. Additionally, since the windows are generated from a randomized ordering of patients, each training epoch exposes the model to different patient groupings, which helps improve generalization and reduces the likelihood of overfitting to specific subsets of the dataset. Overall, the window-based training strategy allows the pipeline to balance computational efficiency with dataset coverage.

## IV. Experiments

### A. Experimental Setup

We evaluated our approach using a subset of 60 patients from the TCGA-PRAD [12] cohort with 79 WSIs. We conducted two experiments: (1) standard joint training and (2) the proposed window-based training. The first used a traditional epoch-based scheme in which the entire training dataset was processed sequentially across epochs. The second applied the window-based strategy, where smaller sequential patient subsets were processed in windows. Timing measurements were recorded for key pipeline components, including data loading, feature extraction, attention-based aggregation, and total inference time. We considered an independent test set of 154 patients from TCGA-PRAD for inference and evaluation, with no overlap with the training set. All experiments were implemented in Python 3.10 with PyTorch and CUDA acceleration, together with NumPy, Pandas, and OpenSlide, and were conducted on an NVIDIA Tesla G4 GPU in the Google Colab environment.

### B. Model Training Setup

In the standard joint-training setup, all WSIs from the selected 60-patient subset were jointly used to train the model. This baseline was evaluated after 1, 5, and 10 epochs to assess predictive performance and cumulative training cost under conventional slide-level MIL training, with separate models trained and evaluated at each epoch setting. In the proposed window-based training, the same 60 patients subset was partitioned into three sequential windows of 20 patients each. During each stage, all WSIs associated with patients in the active window were used for training and the learned weights are frozen and fine-tuned on subsequent windows. The proposed approach was evaluated under 5 and 10-epoch schedules for the 154 patients in the testing set and across different values of *E*_*w*_, where *E*_*w*_ denotes the number of training epochs applied within each window before advancing to the next one while *E* denotes the epochs in standard joint training approach. We used a relatively larger test set to obtain a more robust and representative estimate of out-of-sample performance. In small-cohort studies, performance estimates can be unstable when the test set is too small; therefore, a larger hold-out cohort was prioritized to enable a more reliable evaluation of model generalizability.

### C. Evaluation Metrics and Efficiency Measurements

We evaluated predictive performance using slide-level accuracy, ROC-AUC, precision, recall, and F1-score. To assess computational efficiency, we computed total training time for each experimental setting and compared in both experimental settings.

## V. Results

We compared the proposed window-based staged training strategy against standard joint training. All reported results correspond to slide-level evaluation on the held-out test set.

### A. Experiment 1: Standard Joint Training

Table I summarizes the performance of conventional joint training across different epoch settings. As the number of training epochs increased, classification performance improved consistently. The best overall results were achieved at *E*=10, yielding an accuracy of 0.9066, ROC-AUC of 0.9427, precision of 0.8912, recall of 0.8844, and F1-score of 0.8878. However, these gains were accompanied by a substantial increase in training time, rising from 04 hour 27 minutes at *E*=1 to 20 hours at *E*=5 and 27 hours 19 minutes at *E*=10 as shown in Table 1.

**TABLE I.** Performance Under Joint Training Across Epoch Settings.

| $E$ | Time (hh:mm:ss) | Acc | AUC | Prec | Rec | F1 |
| --- | --- | --- | --- | --- | --- | --- |
| 1 | 04:27:05 | 0.7602 | 0.8034 | 0.7489 | 0.7321 | 0.7404 |
| 5 | 20:00:53 | 0.8630 | 0.9026 | 0.8564 | 0.8381 | 0.8472 |
| 10 | 27:19:36 | **0.9066** | **0.9427** | 0.8912 | 0.8844 | 0.8878 |

**TABLE II.** Performance Evaluation with a 5-Epochs-Per-Window Training Scheme.

| $E_w$ | Time (hh:mm:ss) | Acc | AUC | Prec | Rec | F1 |
| --- | --- | --- | --- | --- | --- | --- |
| 1 | 02:44:00 | 0.7291 | 0.7809 | 0.7089 | 0.7434 | 0.7256 |
| 2 | 03:33:00 | 0.7562 | 0.8200 | 0.7499 | 0.7607 | 0.7553 |
| 3 | 02:10:00 | 0.7805 | 0.8580 | 0.7848 | 0.7912 | 0.7880 |
| **Total** | **08:27:00** | — | — | — | — | — |

**TABLE III.** Performance Evaluation with a 10-Epochs-Per-Window Training Scheme.

| $E_w$ | Time (hh:mm:ss) | Acc | AUC | Prec | Rec | F1 |
| --- | --- | --- | --- | --- | --- | --- |
| 1 | 05:26:36 | 0.7861 | 0.8554 | 0.7824 | 0.7939 | 0.7856 |
| 2 | 05:47:53 | 0.8224 | 0.8678 | 0.8221 | 0.8240 | 0.8230 |
| 3 | 04:41:45 | 0.8397 | 0.8886 | 0.8452 | 0.8436 | 0.8444 |
| **Total** | **15:56:14** | — | — | — | — | — |

### B. Experiment 2: Window-Based Training

Tables II and III present the results of the proposed window-based training strategy under 5-epochs-per-window and 10-epochs-per-window settings, respectively. For the 5-epochsper-window configuration, performance improved progressively with increasing window index, with the best performance observed at *E*_*w*_=3, achieving an accuracy of 0.7805 and F1-score of 0.7880. The total training time for this configuration was 08 hours 27 minutes. Similarly, under the 10-epochs-per-window configuration, the strongest results were also obtained at *E*_*w*_=3, reaching an accuracy of 0.8397, ROC-AUC of 0.8886, precision of 0.8452, recall of 0.8436, and F1-score of 0.8444, with a total training time of 15 hours and 56 minutes.

Overall, the proposed window-based strategy offered a more computationally efficient alternative to conventional joint training (see Figure. 2). While joint training achieved the highest classification performance, the window-based method markedly reduced training time while preserving competitive accuracy. The standard joint training approach in Experiment 1 achieves higher predictive accuracy by repeatedly exposing the model to the entire dataset, while the window-based approach in Experiment 2 improves scalability and system efficiency by reducing data loading overhead. This tradeoff suggests that the proposed approach is particularly well suited for WSI classification settings where computational efficiency is a critical practical constraint.

**Fig. 2.**
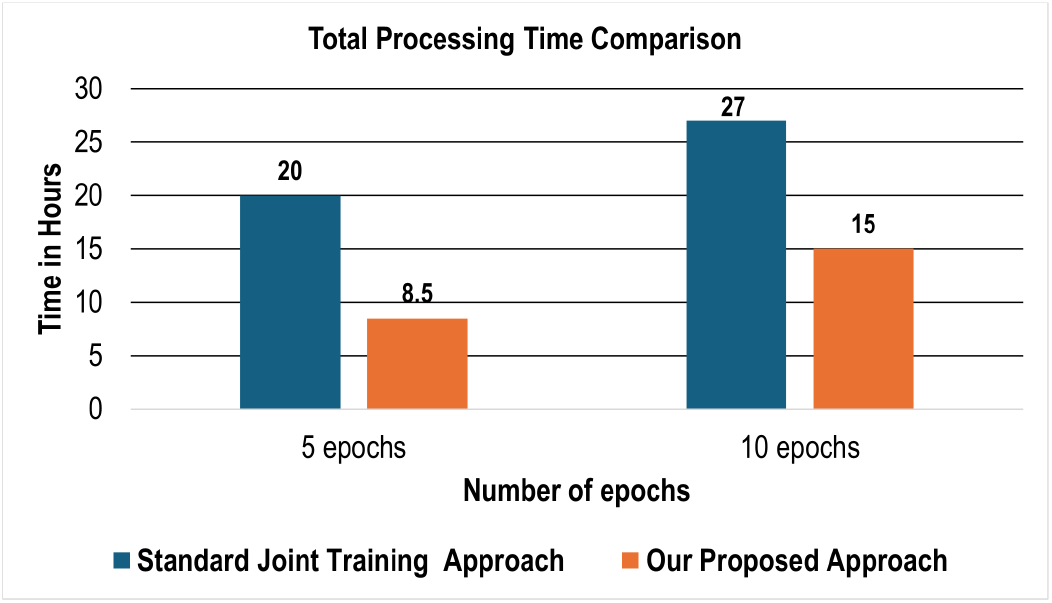
Comparison of total training time across joint training and the proposed window-based training strategy.

## VI. Conclusion

In this paper, we presented an efficient and interpretable learning framework for large-scale histopathology analysis. By combining UNI-based patch encoding, CLAM-SB attention-based multiple instance learning, and a window-based staged training strategy, the proposed approach reduces computational overhead while maintaining competitive slide-level predictive performance. Experiments on TCGA-PRAD show that staged training over patient-grouped windows provides a practical balance between predictive performance and computational efficiency. These findings suggest that window-based optimization is a promising direction for scalable WSI analysis in resource-constrained settings.

## Data Availability

All data produced in the present study are available upon reasonable request to the authors

